# The interactive effect of intra-beat and inter-beat blood pressure variability on neurodegeneration in older adults

**DOI:** 10.1101/2024.05.01.24306724

**Authors:** Trevor Lohman, Fatemah Shenasa, Isabel Sible, Arunima Kapoor, Allison C Engstrom, Shubir Dutt, Elizabeth Head, Lorena Sordo, John Paul M. Alitin, Aimee Gaubert, Amy Nguyen, Daniel A Nation

## Abstract

Blood pressure variability (BPV) and arterial stiffness are age-related hemodynamic risk factors for neurodegenerative disease, but it remains unclear whether they exert independent or interactive effects on brain health. When combined with high inter-beat BPV, increased intra-beat BPV indicative of arterial stiffness could convey greater pressure wave fluctuations deeper into the cerebrovasculature, exacerbating neurodegeneration. This interactive effect was studied in older adults using multiple markers of neurodegeneration, including medial temporal lobe (MTL) volume, plasma neurofilament light (NfL) and glial fibrillary acidic protein (GFAP). Older adults (N=105) without major neurological or systemic disease were recruited and underwent brain MRI and continuous BP monitoring to quantify inter-beat BPV through systolic average real variability (ARV) and intra-beat variability through arterial stiffness index (ASI). Plasma NfL and GFAP were assessed. The interactive effect of ARV and ASI on MTL atrophy, plasma NfL, and GFAP was studied using hierarchical linear regression. Voxel-based morphometry (VBM) was used to confirm region-of-interest analysis findings. The interaction between higher ARV and higher ASI was significantly associated with left-sided MTL atrophy in both the region-of-interest and false discovery rate-corrected VBM analysis. The interactive effect was also significantly associated with increased plasma NfL, but not GFAP. The interaction between higher ARV and higher ASI is independently associated with increased neurodegenerative markers, including MTL atrophy and plasma NfL, in independently living older adults. Findings could suggest the increased risk for neurodegeneration associated with higher inter-beat BPV may be compounded by increased intra-beat variability due to arterial stiffness.

## Introduction

Increased blood pressure variability (BPV) has emerged as a risk factor for stroke ^1^, cerebral small vessel disease (CSVD) ^2,3^, neurodegenerative disease ^4^ and dementia ^5^, independent of average blood pressure. Various markers of BPV have been investigated in the literature including both visit-to-visit BPV and beat-to-beat BPV, with beat-to-beat BPV emerging as a valid ^6,7^, high temporal resolution hemodynamic marker associated with cardiovascular function ^8^, autonomic function ^9^, and vascular aging ^10^. Average real variability (ARV) derived from beat-to-beat blood pressure data is a reliable measure of inter-beat blood pressure variability that is less susceptible to outliers than other BPV metrics ^11–13^. Prior studies have variously quantified ARV and arterial stiffness and identified them separately as risk factors for age-related cognitive decline ^14,15^, dementia risk ^16,17^, and cerebral small vessel disease ^18–20^. The mechanisms underlying the adverse independent effects of increased inter-beat BPV on cerebrovascular and neurodegenerative disease remain unclear, but may be related to the ability of the cerebrovasculature to adapt to chronic fluctuations in pressure and flow while sustaining cerebral perfusion ^21^, blood-brain barrier integrity ^22^ and clearance of brain waste products ^23^.

Arterial stiffening has also been implicated in cerebrovascular ^24,25^ and neurodegenerative disease ^17,26^. Arterial stiffness entails age-related loss of compliance and elasticity within the large conducting arteries ^27^ (e.g., aorta and carotids), resulting in the loss of the buffering of pressure wave amplitude (i.e., the Windkessel effect ^28^). The loss of pressure wave buffering by conducting arteries leads to transmission of higher amplitude pressure waves into muscular and resistance arteries, contributing to cerebrovascular disease and disrupting cerebral microvascular function ^24,29^. Arterial stiffness also leads to uncoupling of systolic and diastolic blood pressure ^30^, resulting in increased intra-beat BPV (i.e., systolic vs. diastolic pressure). Intra-beat BPV can be quantified during continuous blood pressure monitoring by the correlation between systolic and diastolic pressure within each beat. This arterial stiffness index (ASI) has been studied extensively over 24-hour time periods (ambulatory arterial stiffness index) ^31,32^, but can also be quantified over shorter time periods. ASI is correlated with measures of arterial stiffness including pulse wave velocity ^33^, pulse pressure ^34^, as well as atherosclerosis ^35^, left ventricular mass index/relative wall thickness ^34^, and can differentiate between young and old vasculature ^36^.

Markers of increased intra-beat BPV, such as ASI and pulse pressure, have been studied as stiffness-related hemodynamic markers of interest that, like inter-beat BPV, are correlated with cerebrovascular disease and neurodegenerative disease ^26,37^. While inter-beat and intra-beat variability are related hemodynamic markers, they are physiologically distinct aspects of blood pressure fluctuation as illustrated conceptually in **Figure 1a-1b** that exist to varying degrees among individuals. Arterial stiffening, indexed by higher intra-beat BPV, promotes transmission of high amplitude pressure waves deeper into the cerebral microvasculature. Thus, higher inter-beat BPV in the presence of arterial stiffness may promote transmission of even greater blood pressure fluctuations, both within (intra-beat) and between (inter-beat) cardiac cycles. The transmission of these fluctuating pressure waves deep into the brain could promote brain injury and neurodegeneration by interfering with cerebral perfusion ^21^, blood brain barrier integrity ^22^, and waste clearance ^23,38^ mechanisms (**Figure 2**).

**Figure 1:**
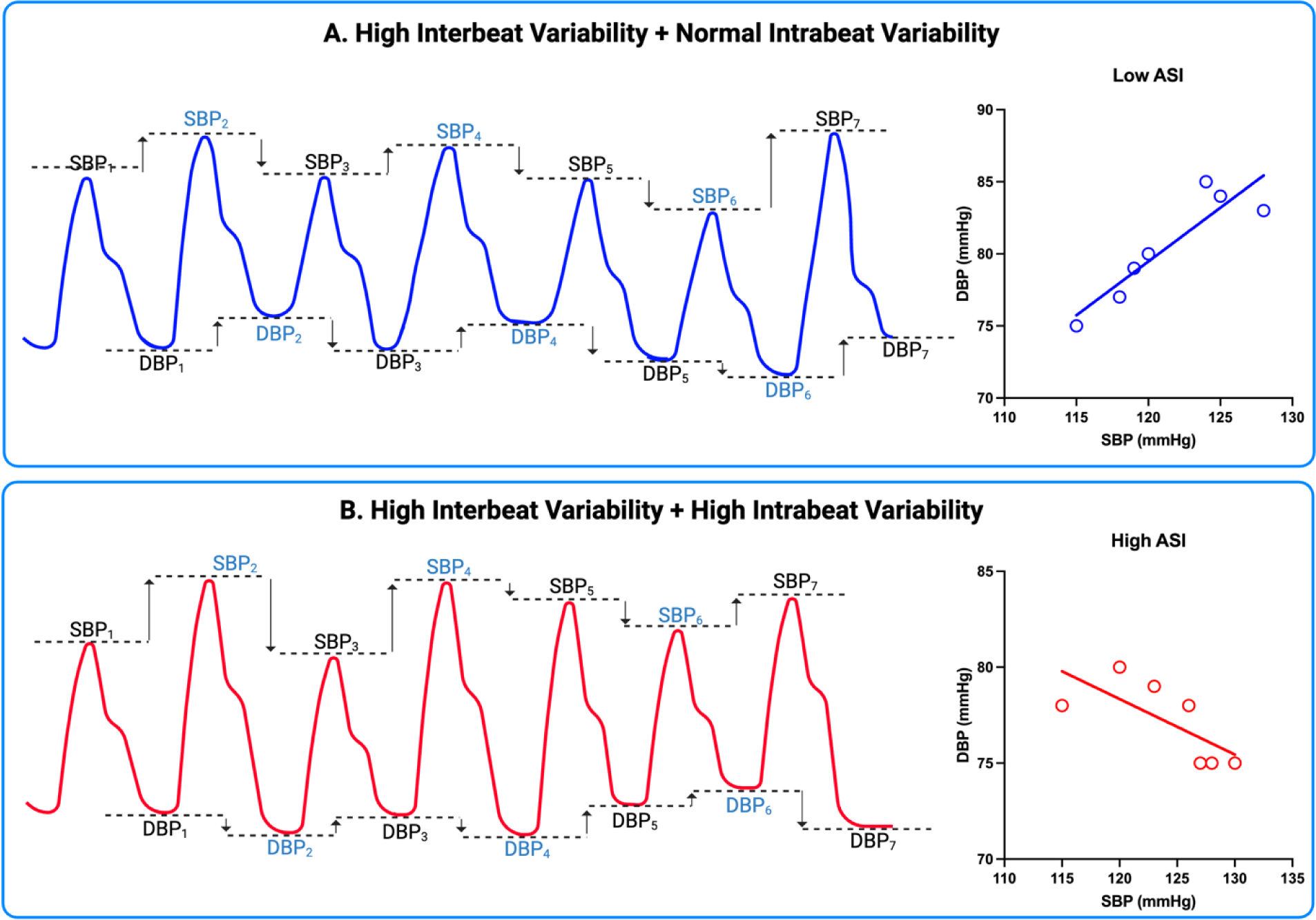
Inter-beat and Intra-beat blood pressure variability. **A:** conceptual continuous blood pressure waveform illustrating high inter-beat blood pressure variability and minimal intra-beat blood pressure variability. The correlation between each cardiac cycle’s systolic blood pressure (SBP) peak and diastolic blood pressure (DBP) trough is illustrated, indicating a positive correlation, indicative of a low arterial stiffness index (ASI= 1 - Pearson’s r). **B:** conceptual continuous blood pressure waveform illustrating high inter-beat blood pressure variability combined with high intra-beat blood pressure variability. The correlation between each cardiac cycle’s SBP peak and DBP trough is illustrated showing an inverse correlation, which is indicative of a high arterial stiffness index (ASI= 1 - Pearson’s r).

**Figure 2:**
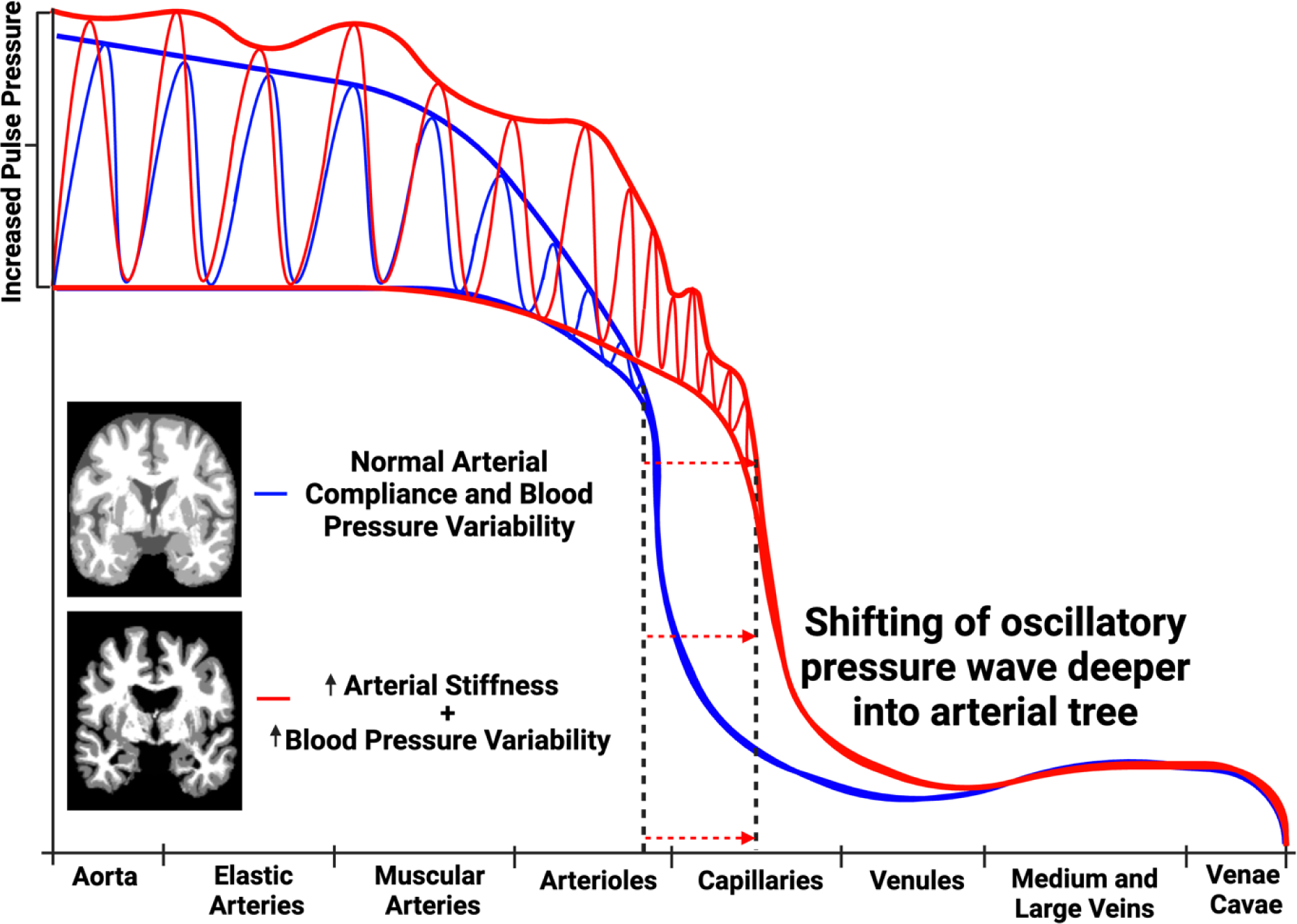
Fluctuating pulse wave transmission hypothesis. An illustration of two arterial pulse waveforms as they transit through systemic circulation. With increased BPV and arterial stiffening there is both greater fluctuation in the oscillation of the pulse wave and greater transmission of the pulse wave oscillations into the cerebral microvascular compartment. Arterial pressure is indicated on the y-axis and the x-axis indicates sequential portions of the arterial tree.

To our knowledge, no studies to date have addressed the potential interaction between inter-beat BPV (ARV) and intra-beat BPV (ASI) relating to neurodegeneration markers on MRI (e.g., MTL atrophy) and in plasma [e.g., neurofilament light (NfL) ^39^ and glial fibrillary acidic protein (GFAP)] ^40^. The MTL has been previously identified as susceptible to both age-related hemodynamic insults and neurodegeneration ^41^, thus the authors hypothesize that the MTL will be differentially affected by this synergistic interaction, manifesting in MTL volume loss and increased plasma markers of neurodegeneration. To test the hypothesis, the present study will investigate the interactive effect of ARV and ASI on MTL atrophy, plasma NfL, and plasma GFAP. MTL atrophy will be quantified using brain MRI.

## Methods

### Participants

Participants were recruited from Orange County communities through outreach events, mailing lists, word-of-mouth, online portals, and other modes of community outreach facilitated by the University of California Irvine (UCI), Alzheimer’s Disease Research Center. All procedures were conducted as part of the Vascular Senescence and Cognition (VaSC) Study at UCI. All participants were independently living at the time of recruitment and were aged 55 to 89 years. Study exclusions were a prior diagnosis of dementia, history of clinical stroke, family history of dominantly inherited neurodegenerative disorders, current neurological or major psychiatric disorders that may impact cognitive function, history of moderate-to-severe traumatic brain injury, current use of medications impairing the central nervous system, current organ failure or other uncontrolled systemic illness, and contraindications for brain MRI. Eligibility for the study was verified via clinical interview and review of current medication with both the participant, and an informant study partner when available. The study was approved by the UCI Institutional Review Board (HS-2019-5324), and all participants gave informed consent.

### Continuous BP Data Acquisition

Participants were asked to take medications as normally prescribed and abstain from caffeine the morning of data collection. Beat-to-beat BP measurements were obtained continuously during supine rest during brain MRI, using an MRI compatible non-invasive continuous BP finger cuff device (Biopac®). First, the participant rests for 3 minutes in the supine position prior to the calibration period. During calibration, two sequential static pressures are taken during continuous BP waveform acquisition using a calibrated, MRI compatible, automatic BP device with an inflatable brachial artery cuff (TeslaDUO). These static pressures are used to calibrate the continuous BP monitor using the Caretaker® system (Biopac®). After calibration, continuous BP was monitored during MRI for 7 minutes and further data processing was performed as previously described ^42^.

### Systolic Blood Pressure Average Real Variability

ARV calculates the average of absolute changes between consecutive blood pressure readings and is calculated as:

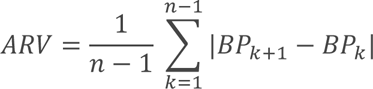

Where n represents the number of blood pressure readings obtained during continuous blood pressure monitoring and k represents the beat index of the readings as previously described^11,42^.

### Arterial Stiffness Index

ASI ^43^ is an indirect measure of arterial stiffness that is correlated with pulse wave velocity ^33^, and atherosclerosis ^35^. ASI was calculated as 1 minus the regression slope of diastolic blood pressure on SBP obtained during 7-minutes of continuous BP monitoring. Higher correlation between systolic and diastolic BP reflects increased arterial compliance, thus a higher ASI (1-Pearson’s r) reflects increased arterial stiffness.

### Plasma Markers of Neurodegeneration

Participant blood samples were obtained by venipuncture and plasma was separated by centrifugation. All plasma samples were kept frozen at -80°C until assay. Plasma levels of GFAP and NfL were determined using single molecule array (Simoa®) Neurology 2-Plex B (N2PB) Kit (Quanterix), following the manufacturer’s protocol on the HD-X machine. Accepted ranges were as follows: NfL = 0 – ∼2000 pg/mL and GFAP = 0 – ∼40000 pg/mL. All biomarker assays were conducted in the same lab at UCI (EH).

### Brain Volume

All participants underwent brain MRI scans conducted on a 3T Siemens Prisma scanner with 20-channel head coil. High-resolution 3D T1-weighted anatomical (Scan parameters: TR = 2300 ms; TE = 2.98 ms; TI = 900 ms; flip angle = 9 deg; FOV = 256 mm; resolution = 1.0 × 1.0 × 1.2 mm^3^; Scan time = 9 min) images were acquired, using 3-dimensional magnetization-prepared rapid gradient-echo (MPRAGE) sequences.

For region-of-interest analysis, post-processing of MPRAGE scans included FreeSurfer 5.3 semi-automated segmentation and parcellation algorithm for quantification of bilateral hippocampal volume (FreeSurfer 6.0) and entorhinal cortical thickness. After automated segmentation, each individual subject was checked for any inaccuracies or misclassifications; manual corrections were made as needed with FreeSurfer’s built-in editing tools, cases were then re-processed, and resulting volumes were used for analyses. For voxel-level indexing of brain regional volume by voxel-based morphometry (VBM) analysis, post-processing was conducted using SPM12 ^44^ and CAT12 ^45^. T1-weighted images were segmented into gray and white matter tissue classes using SPM12’s unified segmentation procedure, spatially normalized, and smoothed with an 8 mm full-width at half-maximum (FWHM) isotropic Gaussian kernel; the resulting gray matter images were checked for sample homogeneity to identify the presence of any potential outliers prior to VBM analysis.

### Vascular Risk Factors

Vascular risk factor (VRF) burden was determined through clinical interviews with the participant and informant (when available), and review of current medications and medical history. The assessed VRFs included history of cardiovascular disease (e.g., heart failure, angina, stent placement, coronary artery bypass graft, intermittent claudication), hypertension, hyperlipidemia, type 2 diabetes, atrial fibrillation, left ventricular hypertrophy and transient ischemic attack. Total VRFs were summed for each participant and elevated VRF burden was defined as ≥2 VRFs (vs. 0-1) as described previously ^46,47^.

## Data Analysis

Data from 105 community-dwelling older adults characterized by demographics, brain MRI, and beat-to-beat ARV were screened for outliers (±3 SD). One participant with total intracranial volume (TIV) normalized left hippocampal volume 4.35 SD above the mean and right hippocampal volume 4.31 SD above the mean was removed from the ROI analysis. No outliers were identified during the VBM sample homogeneity screen. A subset of these participants (n=56) also had plasma NfL and GFAP levels for statistical analysis.

Hierarchical linear regression models assessed the statistical significance of the relationship between the ARV*ASI interaction term and both MTL atrophy and plasma markers of neurodegeneration adjusted for age, sex, ARV, and ASI. MTL atrophy analyses also included TIV as a covariate. For all models, step 0 included age, sex, and SBP, step 1 added ARV and ASI, and step 2 added the ARV*ASI interaction term. A multicollinearity screen was performed for all predictor variables, with a variance inflation factor (VIF) above 4 indicating significant multicollinearity. VIF was <2 for all predictors used in the present analyses. All analyses were performed in R ^48^, with conditional effects figures generated using the Hayes PROCESS macro ^49^ model 1 (simple moderation) in R.

For VBM analyses, voxel-wise multiple regression was conducted with the ARV*ASI product term as the independent variable, gray matter map as the dependent variable, and total intracranial volume as a covariate. As this was a confirmatory analysis based on ROI analysis results, we corrected for false discovery rate and family-wise error but also conducted analyses with the uncorrected significance threshold set at *p* < .001.

## Results

Participant characteristics and their demographics are displayed in **Table 1**. Higher ARV was associated with increased age (*B=* 3.43, *P*= .00002) and with higher ASI (*B=* .08, *P=* .0000004). Higher ASI trended towards a significant relationship with increased age (*B*= 9.90, *P*= .05). No sex differences were observed for either variable.

**Table 1:** Participant Characteristics and Demographics (n=105)

| Variable Name | Mean±SD |
| --- | --- |
| Age | 69.52±6.67 |
| Sex (n, female%) | 66 (62.86) |
| Race/ethnicity |  |
| Asian (n, %) | 19 (18.10) |
| Native Hawaiian or Pacific Islander (n, %) | 0 (0) |
| American Indian or Alaska native (n, %) | 0 (0) |
| Black or African American (n, %) | 1 (.95) |
| White (n, %) | 83 (79.05) |
| Other (n, %) | 2 (1.90) |
| ≥2 Vascular Risk Factors (n, %) | 47 (44.76) |
| Hypertension (n, %) | 37 (35.23) |
| High cholesterol (n, %) | 52 (49.52) |
| Diabetes (n, %) | 11 (10.48) |
| Current smoker (n, %) | 33 (31.43) |
| Cardiovascular disease (n, %) | 11 (10.48) |
| Atrial fibrillation (n, %) | 5 (4.76) |
| Transient ischemic attack (n, %) | 2 (1.90) |
| Left hippocampal volume (mm<sup>3</sup>) (n=104) | 3812.54±466.16 |
| Right hippocampal volume (mm<sup>3</sup>) (n=104) | 3975.62±364.81 |
| Left entorhinal cortex thickness (mm) (n=104) | 3.17±.33 |
| Right entorhinal cortex thickness (mm) (n=104) | 3.34±.33 |
| Plasma NfL (pg/ml) (n=56) | 18.86±8.31 |
| Plasma GFAP (pg/ml) (n=56) | 148.44±73.943 |
| ARV (mmHg) | 1.68±.79 |
| ASI (1-Pearson's r) | .60±.13 |
| Average SBP | 133.00±16.95 |

NfL: neurofilament light, GFAP: glial fibrillary acidic protein, ARV: average real variability, ASI: arterial stiffness index, Adj: adjusted, pg/ml: picograms per milliliter, SBP: systolic blood pressure, B: unstandardized beta coefficient, SE: standard error, P: p-value.

The ARV*ASI interaction term was significantly associated with decreased left and right hippocampal volumes, independent of age, sex, average SBP and TIV (**Table 2a-2b**), The adverse effect of higher ARV on hippocampal volume was greatest in participants with higher ASI (left *B=* -252.79, *P=* .0002; right *B=* -193.56, *P=* .001) as shown in **Figure 3a-3b**. ARV*ASI was associated with decreased left entorhinal cortical thickness after accounting for age, sex, average SBP, and TIV (**Table 2c**), and the effect was greatest in participants with high ASI (*B=* - .13, *P=* .007) as shown in **Figure 3c**. No similar significant relationship between the ARV*ASI interaction term and right entorhinal cortex thickness was observed (**Table 2d**).

**Table 2:**
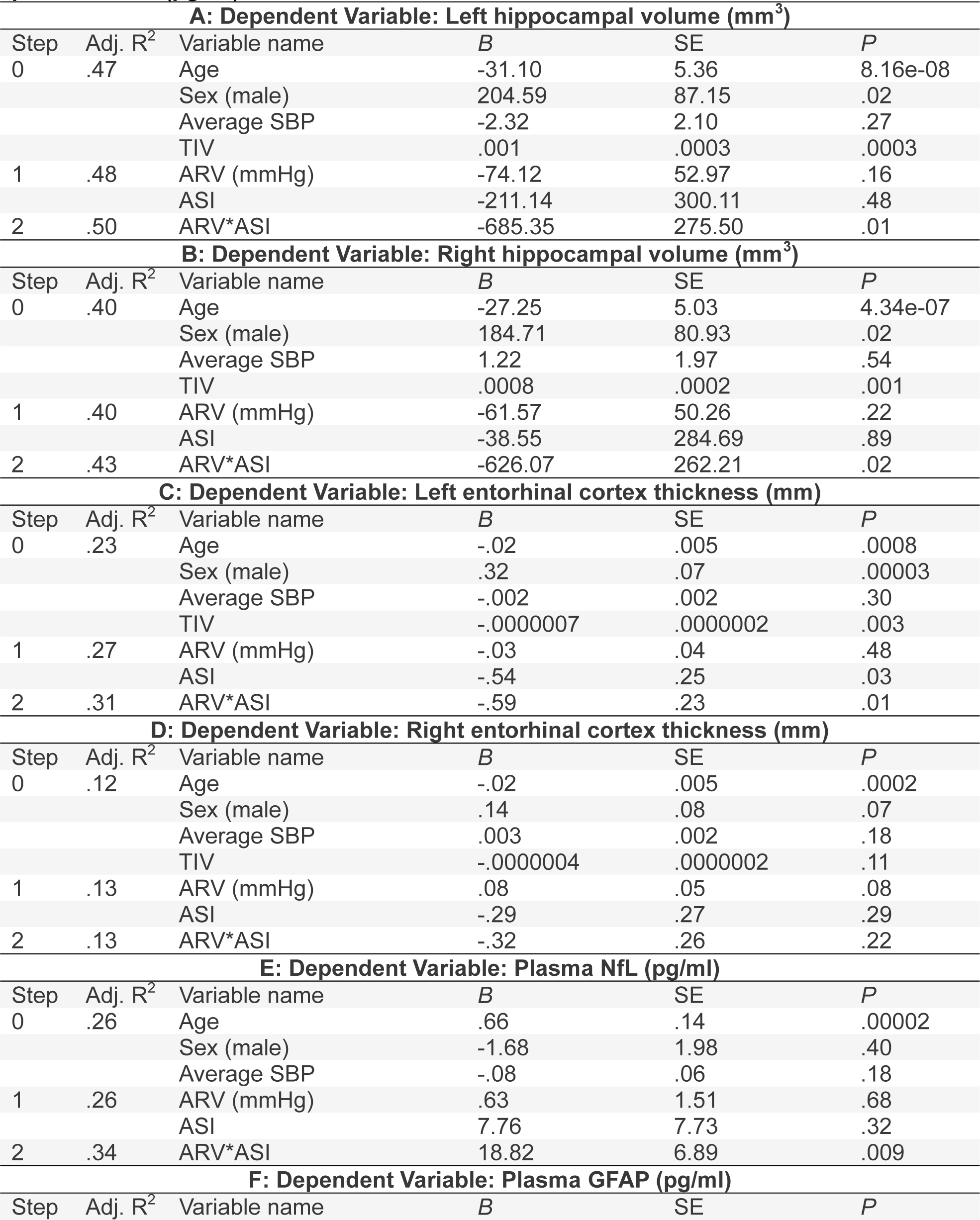
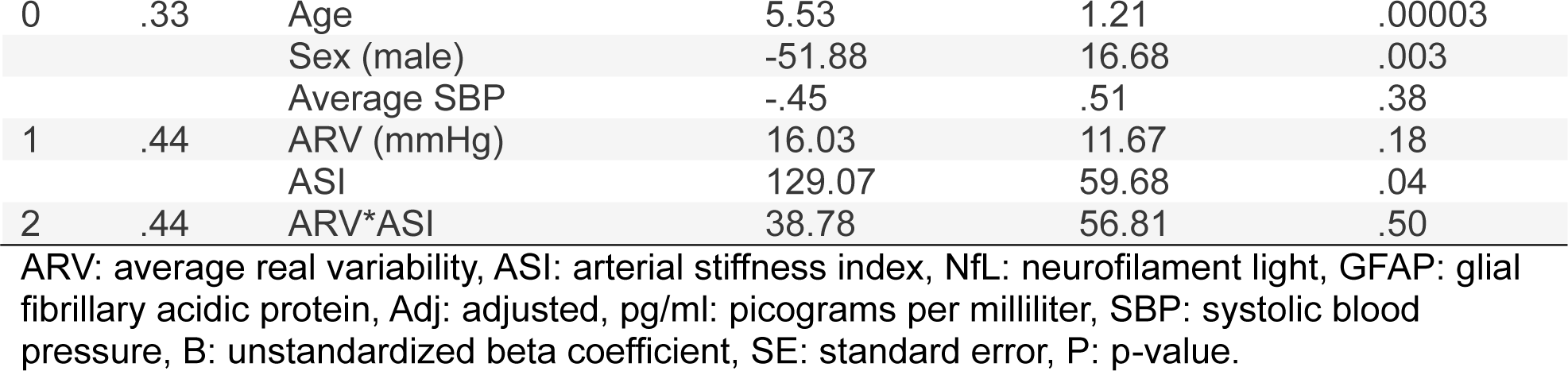
Hierarchical linear regression model parameters assessing the interactive effects of ARV and ASI on A: left hippocampal volume, B: right hippocampal volume, C: left entorhinal cortex thickness, D: right entorhinal cortex thickness, E: plasma NfL, F: plasma GFAP (pg/ml).

**Figure 3:**
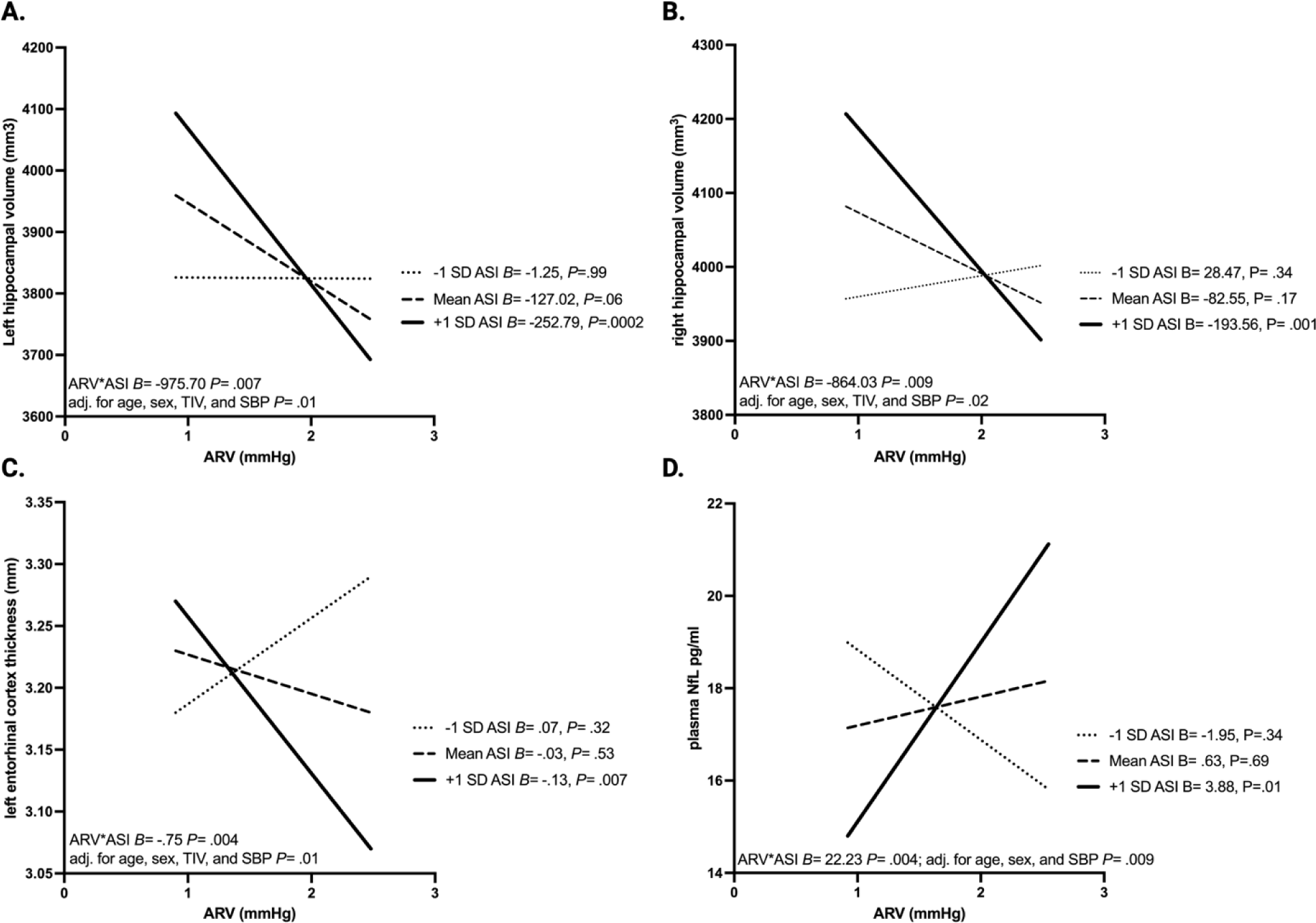
The interaction between ARV and ASI in relation to hippocampal volume, entorhinal cortex thickness, and plasma neurofilament light (NfL). **A.** ARV in mmHg displayed on x-axis and left hippocampal volume (mm^3^) displayed on y-axis. Three groups are displayed (-1 SD ASI, mean ASI, and +1 SD ASI) with unstandardized beta coefficients and p-values for the within group relationship between the BPV – left hippocampal volume relationship. ARV*ASI interaction term unstandardized regression coefficient and p-values displayed above the x-axis. **B.** ARV in mmHg displayed on x-axis and right hippocampal volume (mm^3^) displayed on y-axis. Three groups are displayed (-1 SD ASI, mean ASI, and +1 SD ASI) with unstandardized beta coefficients and p-values for the within group relationship between the BPV – right hippocampal volume relationship. ARV*ASI interaction term unstandardized regression coefficient and p-values displayed above the x-axis. **C.** ARV in mmHg displayed on x-axis and left entorhinal cortex thickness (mm) displayed on y-axis. Three groups are displayed (-1 SD ASI, mean ASI, and +1 SD ASI) with unstandardized beta coefficients and p-values for the within group relationship between the ARV – left entorhinal thickness relationship. ARV*ASI interaction term unstandardized regression coefficient and p-values displayed above the x-axis. **D.** BPV in mmHg displayed on x-axis and plasma NfL in pg/ml displayed on y-axis. Three groups are displayed (-1 SD ASI, mean ASI, and +1 SD ASI) with unstandardized beta coefficients and p-values for the within group relationship between the ARV – plasma NfL relationship. ARV*ASI interaction term unstandardized regression coefficient and p-values displayed above the x-axis.

The ARV*ASI interaction term was also significantly associated with increased plasma NfL (**Table 2e**) but not plasma GFAP (**Table 2f**) adjusting for age, sex, and average SBP. The adverse effect of ARV on plasma NfL was greatest in participants with high ASI (*B=* 3.88, *P=* .01) as shown in **Figure 3d**.

### Voxel-Based Morphometry Analysis

In confirmatory VBM analysis the ARV*ASI interaction term was significantly (uncorrected and FDR corrected) associated with decreased gray matter volume in a cluster centered within the left entorhinal region, left parahippocampal gyrus, left fusiform gyrus, left temporal pole, left inferior temporal gyrus, left amygdala, and left hippocampus (*p*_uncorrected_= .007, *q*_FDR-corrected_= .03, *p*_FWE-corrected_= .10). A second cluster centered around the right fusiform gyrus, right hippocampus, right parahippocampal gyrus, right amygdala, right entorhinal area, and the right inferior temporal gyrus was significant in the peak level analysis (*p*_uncorrected_ <.001) but failed to achieve significance in the cluster level analysis (*p*_uncorrected_ =.55) . VBM results are shown in **Figure 4**.

**Figure 4:**
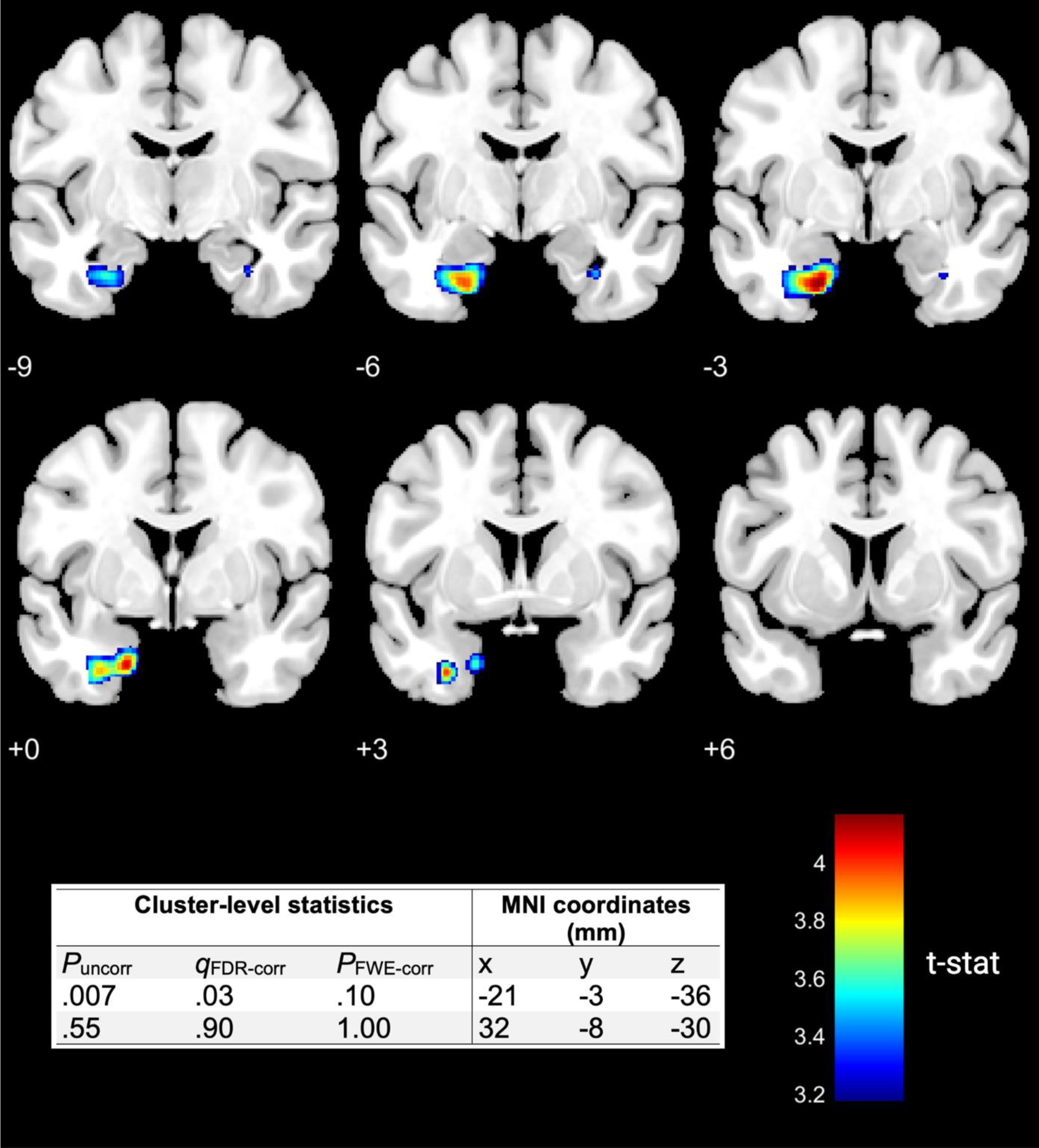
Voxel-based Morphometry. Visualization of gray matter reductions associated with the blood pressure variability*arterial stiffness interaction term. One statistically significant cluster was identified centered around the left entorhinal area, left parahippocampal gyrus, left fusiform gyrus, left temporal pole, left inferior temporal gyrus, left amygdala, and left hippocampus. Cluster level statistics with uncorrected p-value, FDR corrected q-value, and family-wise error p-value displayed alongside a t-statistic color bar with higher t-statistic (t-stat) indicating greater gray matter reduction. MNI coordinates along the y-axis indicated for each coronal slice.

## Discussion

The interaction between increased inter-beat BPV, indexed by ARV, and intra-beat BPV, indexed by ASI, was significantly associated with medial temporal atrophy and increased plasma NfL independent of age, sex, and average SBP. Specifically, adverse effects of increased inter-beat BPV were most apparent in individuals with elevated intra-beat BPV. The ROI volumetric analysis was supported by a VBM analysis which confirmed that the interaction between elevated BPV and ASI was specifically related to medial temporal atrophy. Although causality cannot be inferred from this observational study, these statistical findings are consistent with our *a priori* hypothesis that the combined effect of two different dimensions of BPV, including both inter-beat and intra-beat variation, would have a synergistic effect on neurodegeneration. This hypothesis is based on the theory proposed herein that the combination of age-related arterial stiffness and increased blood pressure fluctuation leads to transmission of fluctuating pulse waves into the cerebral microvasculature causing neurodegeneration. The findings are also consistent with prior studies separately relating visit-to-visit BPV and arterial stiffness to medial temporal perfusion decline and neurodegeneration^21,50–52^.

Both the ROI and VBM analysis revealed increased left-sided involvement, which is consistent with known left-sided neurodegenerative susceptibility ^53^ in a variety of diseases including Alzheimer’s disease (AD), Parkinson’s disease, frontotemporal dementia, and Huntington’s disease. Interestingly, atherosclerotic plaque development and intima-media common carotid thickening also show a left-sided predilection ^54^, potentially due to increased left-sided hemodynamic stress ^54^. Left-sided cerebrovascular events may also be more common than right-sided events ^55,56^. The finding that the interaction between two hemodynamic markers was similarly associated with left-sided MTL atrophy and neurodegenerative plasma markers in the present study could suggest a shared vascular mechanism for neurodegeneration.

Plasma NfL, but not GFAP, was associated with the ARV-ASI interaction, such that greater ARV in the presence of high ASI correlated with higher plasma NfL levels. Plasma NfL is more sensitive than GFAP to the presence of CSVD ^57^, and both BPV and arterial stiffness are associated with CSVD, potentially accounting for the findings of the present study ^2,58^. Also, plasma NfL reflects the current severity of neurodegeneration based on MTL atrophy, hypometabolism, and white matter integrity loss ^39^ , while plasma GFAP is only indirectly associated with neuronal injury, being a marker of the reactive astrogliosis that can occur during neurodegeneration ^59^. Given the present study findings relating the ARV-ASI interaction to be both MTL volume levels and plasma NfL levels, the combined effect of BPV and arterial stiffness may be related to both vascular brain injury and neurodegeneration. Although MTL atrophy indicative of neurodegeneration is a key feature of the AD “ATN” (N=neurodegeneration) criteria ^60^, neither MTL atrophy nor NfL are disease-specific biomarkers ^61^. Both are more generally associated with a variety of age-related neurodegenerative and neurovascular diseases, including CSVD and vascular dementia ^61,62^, Lewy bodies dementia ^63^ and Parkinson’s disease ^63^. Additionally, both NfL and left-sided MTL volume loss are associated with future cognitive decline ^64,65^, AD ^66^ and progression to dementia ^67^. Similarly, plasma NfL has been identified as a susceptibility marker given its ability to predict future abnormal alterations in brain structure and function ^39^. This could indicate that two hemodynamic markers of vascular aging, BPV and ASI, may interact to confer a susceptibility to future neurological and neurovascular disease in older adults.

Strengths of the present study include the test of a novel hypothesis of the interaction between two hemodynamic risk markers to brain injury and neurodegeneration markers in older adults, the combination of imaging and biofluid markers and the confirmation of ROI-volumetric findings with VBM analysis. Study limitations include the cross-sectional study design which limits causal inference. Further research examining the interaction between inter-beat and intra-beat BPV is needed to fully elucidate the relationship between blood pressure fluctuation and risk of neurodegeneration. The novel findings presented here may have significant clinical implications and offer new insights into the pathogenesis of age-related neurodegeneration.

### Perspectives

The present study proposes and tests the fluctuating pulse wave transmission hypothesis, in which blood pressure fluctuations are transmitted further into the delicate cerebral microvasculature. To accomplish this, the effect of the interaction between an index of arterial stiffness (intra-beat BPV) and beat-to-beat BPV was assessed using plasma and neuroimaging biomarkers of medial temporal neurodegeneration. Participants with higher intra-beat and inter-beat BPV displayed greater MTL atrophy and increased plasma NfL. Future studies should assess the interaction of BPV and pulse wave velocity to determine if the observed effects are specific to the interaction between BPV and arterial stiffening, or if BPV is interacting with another physiological phenomenon captured by intra-beat BPV such as ventricular contractility or pulse pressure variability ^34,68^.

### Novelty and Relevance

1. What Is New?

- The discovery of an interactive effect of inter-beat BPV and intra-beat BPV on neurodegeneration markers representing the combined influence of blood pressure fluctuation and arterial stiffness.
2. What is relevant?

- Higher intra-beat and inter-beat BPV is associated with greater MTL atrophy and increased plasma NfL independent of mean blood pressure level.
3. Clinical/Pathophysiological Implications?

- These initial results indicate that further study of the synergistic effects of BPV and arterial stiffness are needed to better understand potentially modifiable hemodynamic risk factors for neurodegeneration that are independent of currently treated mean blood pressure levels.

## Non-standard Abbreviations and Acronyms

ARV: average real variability
ASI: Arterial stiffness index
BPV: blood pressure variability
GFAP: glial fibrillary acidic protein
MTL: medial temporal lobe
NfL: neurofilament light
SBP: systolic blood pressure
VBM: voxel-based morphometry

## Statements and Declarations

### Funding

This research was supported by National Institutes of Health grants (DAN: R01AG064228, R01AG060049, R01AG082073, P01AG052350, P30AG066530), (EH: UCI ADRC P30AG066519), and the Canadian Institutes of Health Research (AK: DFD-170763).

### Competing Interests

The authors have no competing interests to declare.

### Data availability statement

The anonymous data that support the findings of this study are available upon reasonable request from the corresponding author, DN, through appropriate data sharing protocols.

## Notes

### Competing Interest Statement

The authors have declared no competing interest.

### Author Declarations

The study was approved by the UCI Institutional Review Board (HS-2019-5324), and all participants gave informed consent.

